# Heart rate variability and electrocardiographic parameters among workers exposed to chronic intermittent hypobaric hypoxia in the Peruvian Andes: A cross-sectional study Cardiac autonomic function in Andean workers

**DOI:** 10.64898/2026.10.15.26365599

**Authors:** Elizbet Susan Montes-Madariaga, Jhon Brando Ortiz-Saavedra, Julio Samuel Mamani-Castillo, Anel Ivonne Valencia-Pacheco, Jorge Andres Ballón Echegaray, Irmia Luz Paz Torres, Ricardo Alfredo Julián León-Vásquez

**Author notes:** **Correspondence:** (RJ).

## Abstract

Chronic intermittent hypobaric hypoxia is an occupational exposure experienced by workers who alternate between high-altitude worksites and lower-altitude resting locations. Its effects on cardiac autonomic modulation and electrocardiographic parameters remain insufficiently characterized in Andean workers. This study aimed to compare heart rate variability and electrocardiographic parameters between workers exposed to chronic intermittent hypobaric hypoxia and controls working permanently at lower altitude in Arequipa, Peru.

An observational cross-sectional study was conducted in 96 male workers aged 20–60 years. The exposed group included 53 workers performing rotational shifts at altitudes ≥3,000 m above sea level with rest periods at <2,500 m, while the control group included 43 workers working permanently at <2,500 m. Heart rate variability was assessed during the morning rest period in Arequipa, at approximately 2,300 m above sea level, using a Polar® H10 heart rate sensor after 15 minutes of rest. Participants also underwent standard 12-lead electrocardiography.

Vital signs were clinically similar between groups. No statistically significant differences were observed in PR interval, corrected QT interval, time-domain heart rate variability parameters, or frequency-domain heart rate variability parameters.

In this sample of apparently healthy male workers evaluated during their rest period at moderate altitude, exposure to chronic intermittent hypobaric hypoxia was not associated with significant differences in resting heart rate variability or standard electrocardiographic parameters.

## Introduction

Cardiovascular diseases remain a leading cause of morbidity and mortality worldwide, with elevated blood pressure being one of the most important modifiable risk factors for cardiovascular events and premature death [1]. In Peru, cardiovascular diseases also represent a major public health concern and are among the principal causes of mortality [2,3]. This burden is particularly relevant in occupational settings where workers are repeatedly exposed to environmental and physiological stressors that may modify cardiovascular regulation.

Chronic intermittent hypobaric hypoxia (CIHH) occurs in workers who alternate between high-altitude workplaces and lower-altitude resting locations. This exposure pattern is common in mining, transport, and other high-altitude industries in the Andean region. Unlike permanent high-altitude residence, CIHH involves repeated cycles of ascent, hypoxic exposure, partial acclimatization, descent, and recovery. These cycles may induce adaptive responses, but they may also produce cardiovascular stress, particularly when combined with rotating shifts, altered sleep patterns, physical workload, and other occupational demands [4,5].

Previous studies have reported heterogeneous findings regarding the cardiovascular effects of CIHH. Experimental studies have suggested potentially beneficial effects, including modulation of the renin-angiotensin system, reduction of blood pressure, and protection against myocardial remodeling under certain conditions [6,7]. However, models simulating alternating work at high and low altitude have also reported cardiac dysfunction, ventricular remodeling, reduced antioxidant capacity, and impaired vascular function [8]. In humans, evidence is similarly inconclusive: while some studies suggest adaptive or cardioprotective mechanisms [9,10], others have reported associations between CIHH and increased cardiometabolic risk, altered blood pressure patterns, right ventricular changes, and elevated pulmonary artery pressure in high-altitude workers [11–14].

Heart rate variability (HRV) is a non-invasive marker of cardiac autonomic modulation and reflects the dynamic interaction between sympathetic and parasympathetic influences on heart rate. In high-altitude contexts, HRV has been used to evaluate autonomic adaptation to hypoxia, physical stress, and acclimatization [15–17]. Time-domain indices such as SDNN, RMSSD, and pNN50 provide information on overall variability and vagal modulation, whereas frequency-domain indices such as LF, HF, and the LF/HF ratio are commonly used to explore autonomic regulation, although the interpretation of LF/HF remains controversial [18,19]. Electrocardiographic parameters, including cardiac axis, PR interval, QT interval, and corrected QT interval, may provide additional information on cardiac conduction and repolarization in individuals exposed to altitude-related hypoxia [20–22].

Despite the relevance of these markers, the cardiovascular response of Peruvian workers exposed to CIHH remains insufficiently characterized. Most available evidence comes from other high-altitude occupational contexts, particularly mining populations in Chile or experimental models, and may not be directly generalizable to workers in the Peruvian Andes. Therefore, this study aimed to compare heart rate variability and electrocardiographic parameters between workers exposed to chronic intermittent hypobaric hypoxia through rotational shifts at high altitude and workers performing permanent activities at lower altitude in Arequipa, Peru.

## Materials and Methods

### Ethical considerations

The research protocol was reviewed and approved by the Institutional Research Ethics Committee of the Universidad Peruana Cayetano Heredia under registration code 21005 and certificate number R-028-01-23. All participants provided written informed consent before enrolment. The study was conducted in accordance with the ethical principles of the Declaration of Helsinki [23]. Data confidentiality was ensured through coded identification and restricted access to the study database.

### Study design and setting

An observational, cross-sectional study was conducted in Arequipa, Peru, located at approximately 2,300 m above sea level. Participants were classified into two groups according to occupational exposure to chronic intermittent hypobaric hypoxia. The exposed group included workers performing rotational shifts at high altitude, defined as ≥3,000 m above sea level, with rest periods at lower altitude, defined as <2,500 m above sea level. The control group included workers with permanent work schedules at lower altitude, defined as <2,500 m above sea level.

### Participants

The study population consisted of male workers aged 20 to 60 years. Participants in the exposed group were eligible if they had performed rotational work at high altitude for at least six consecutive months, with rest periods at lower altitude. Participants in the control group were eligible if they worked permanently at lower altitude.

A total of 100 volunteers were initially enrolled. After applying the eligibility criteria, 96 participants were included in the final analysis: 53 workers exposed to chronic intermittent hypobaric hypoxia and 43 controls.

Only male participants were included to reduce biological variability related to sex-specific hormonal and inflammatory profiles and because the available occupational cohort was predominantly male.

### Inclusion and exclusion criteria

Participants were included if they were male workers aged 20–60 years and met the exposure definition for either the exposed or control group.

Participants were excluded if they had a body mass index <18.5 or >40 kg/m^2^, chronic mountain sickness, previous diagnosis of hypertension, coronary heart disease, diabetes mellitus, cancer, chronic obstructive pulmonary disease, or any chronic inflammatory condition. Participants were also excluded if they had an acute illness or had used antibiotics or anti-inflammatory medication within the previous 30 days. Current smokers, defined as those smoking at least one cigarette per day, and former smokers who had quit within the previous year were also excluded.

### Exposure assessment

Exposure to chronic intermittent hypobaric hypoxia was defined as rotational work at altitudes ≥3,000 m above sea level, alternating with rest periods at <2,500 m above sea level, for at least six consecutive months. Workplace altitude was verified using official employment information provided by the workers’ companies, including workplace location and altitude. Control participants worked continuously at <2,500 m above sea level.

### Outcome variables

Heart rate variability and electrocardiographic parameters were considered the main outcome variables.

Heart rate variability was assessed during the workers’ rest period in Arequipa, Peru, at an altitude of approximately 2,300 m above sea level. Measurements were performed in the morning using a Polar® H10 heart rate sensor (Polar Electro, Kempele, Finland), a chest strap device previously validated for HRV assessment against electrocardiographic recordings [24]. Before recording, participants remained at rest for 15 minutes without physical stimulation before or during the measurement. Heart rate recordings lasted approximately 10 minutes.

Data were collected using the Polar Flow platform and subsequently exported to a Microsoft Excel spreadsheet for database organization. The final dataset was then exported to R/RStudio for statistical analysis. No changes were made to the original values obtained from the HRV recordings before statistical processing.

Time-domain HRV parameters included the standard deviation of normal-to-normal intervals, the coefficient of variation of NN intervals, the root mean square of successive differences between adjacent NN intervals, and the percentage of adjacent NN intervals differing by more than 50 ms. Frequency-domain HRV parameters included low-frequency power, high-frequency power, and the LF/HF ratio.

After HRV assessment, all participants underwent a standard 12-lead electrocardiogram. Electrocardiographic parameters included cardiac axis, PR interval, QT interval, and corrected QT interval.

### Statistical analysis

Data collected through Polar Flow were exported to Microsoft Excel for database organization and subsequently analyzed using R/RStudio. R is an open-source environment for statistical computing and graphics [25], and RStudio was used as the integrated development environment for data analysis [26].

Categorical variables were summarized as absolute and relative frequencies. Numerical variables were summarized as medians and interquartile ranges. Between-group comparisons were performed using Student’s t-test or the Mann-Whitney U test, according to the distribution of the data. A p-value <0.05 was considered statistically significant.

## Results

A total of 96 male workers were included in the final analysis. Of these, 53 participants were exposed to chronic intermittent hypobaric hypoxia and 43 were included in the control group. The median age of the overall sample was 34 years. Vital signs were clinically similar between groups, with both groups showing a median systolic blood pressure of 110 mmHg, a median diastolic blood pressure of 70 mmHg, and a median peripheral oxygen saturation of 96%. Heart rate measured by electrocardiogram also showed comparable median values between groups **(Table 1)**.

**Table 1.** Clinical characteristics and vital signs of participants according to chronic intermittent hypobaric hypoxia exposure.

|  | All<br>N = 96 | Control<br>N = 43 | CIHH<br>N = 53 |
| --- | --- | --- | --- |
| Age, years | 34 (27-39) | 34 (25.5-38.0) | 34 (30-39) |
| Pulse, bpm | 70 (62.75-74.00) | 70 (64-75.5) | 70 (62-74) |
| Heart rate by ECG, bpm | 58 (54.00-63.25) | 58 (54.00-64.5) | 58 (54.00-63.00) |
| Respiratory rate,<br>breaths/min | 16 (16-17) | 16 (16-16) | 16 (16-17) |
| Systolic blood pressure,<br>mmHg | 110 (100-115) | 110 (100-112.5) | 110 (100-115) |
| Diastolic blood pressure,<br>mmHg | 70 (60-70) | 70 (60-70) | 70 (60-70) |
| SpO<sub>2</sub>, % | 96 (95-96) | 96 (95-96) | 96 (95-96) |

Data are presented as median and interquartile range. CIHH: chronic intermittent hypobaric hypoxia; ECG: electrocardiogram; SpO<sub>2</sub>: peripheral oxygen saturation.

Electrocardiographic parameters showed similar distributions between workers exposed to chronic intermittent hypobaric hypoxia and controls. PR interval values did not differ significantly between groups (p = 0.879). Likewise, corrected QT interval values were comparable between groups, with no statistically significant difference observed (p = 0.809) **(Fig 1)**.

**Fig 1.**
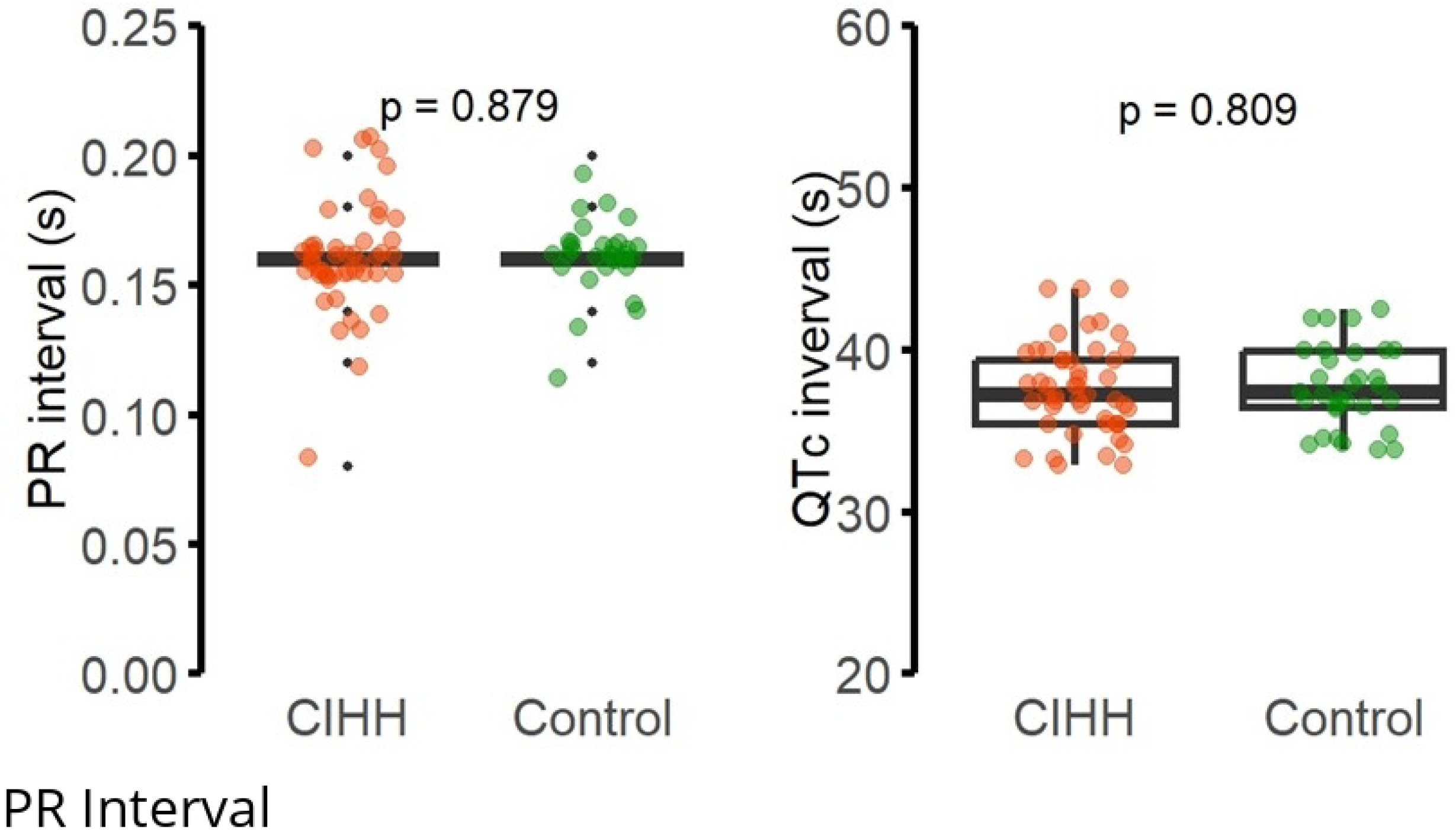
Electrocardiographic parameters according to chronic intermittent hypobaric hypoxia exposure. The figure shows the distribution of PR interval and corrected QT interval among workers exposed to chronic intermittent hypobaric hypoxia and controls. Boxplots represent the median and interquartile range, with individual data points overlaid. No statistically significant differences were observed between groups for PR interval or QTc interval. CIHH: chronic intermittent hypobaric hypoxia; QTc: corrected QT interval. Time-domain heart rate variability parameters also showed no statistically significant differences between groups. Overall variability, assessed through SDNN, was comparable between workers exposed to chronic intermittent hypobaric hypoxia and controls (p = 0.273). Similarly, the coefficient of variation of NN intervals (p = 0.150), RMSSD (p = 0.830), pNN50 (p = 0.735), SEM (p = 0.224), and sample entropy (p = 0.180) showed no significant between-group differences **(Fig 2)**.

**Fig 2.**
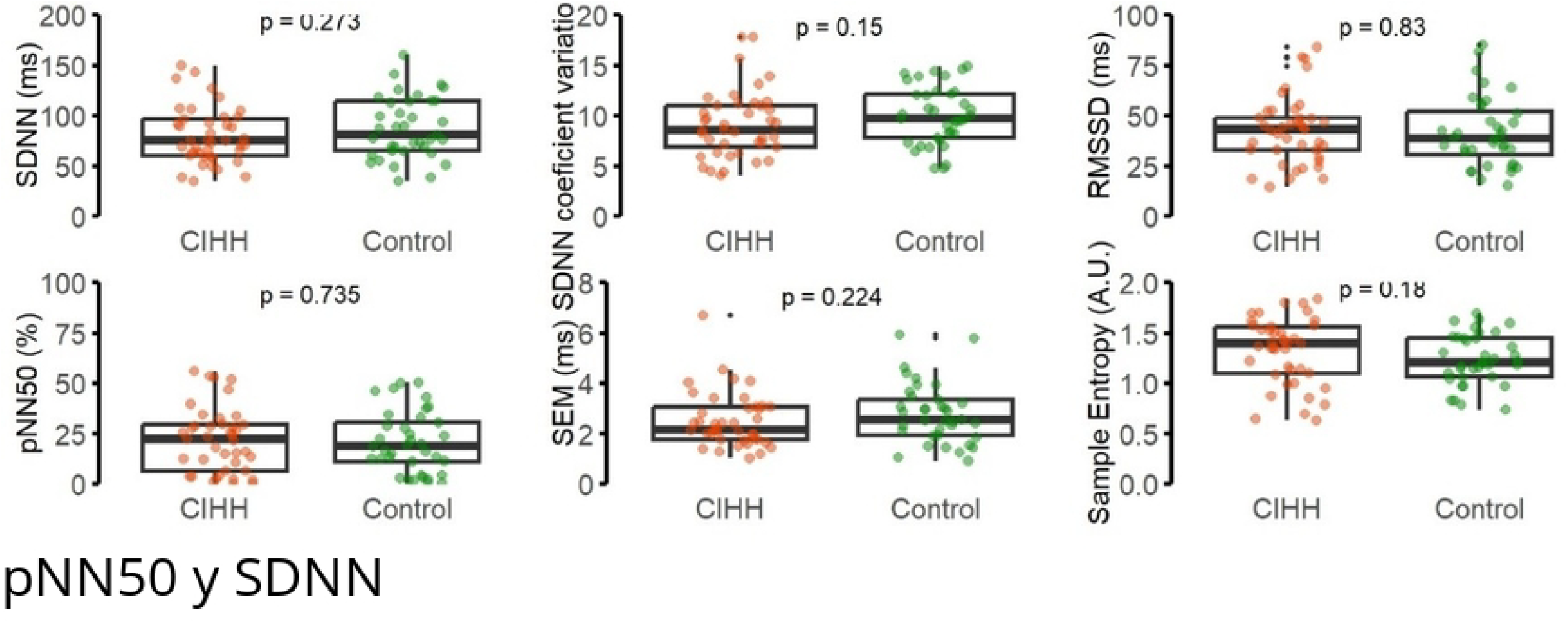
Time-domain heart rate variability parameters according to chronic intermittent hypobaric hypoxia exposure. The figure shows the distribution of time-domain heart rate variability parameters among workers exposed to chronic intermittent hypobaric hypoxia and controls. Parameters included standard deviation of normal-to-normal intervals, coefficient of variation of NN intervals, root mean square of successive differences between adjacent NN intervals, percentage of adjacent NN intervals differing by more than 50 ms, standard error of the mean, and sample entropy. Boxplots represent the median and interquartile range, with individual data points overlaid. No statistically significant differences were observed between groups. CIHH: chronic intermittent hypobaric hypoxia; SDNN: standard deviation of NN intervals; RMSSD: root mean square of successive differences; pNN50: percentage of adjacent NN intervals differing by more than 50 ms; SEM: standard error of the mean. Frequency-domain heart rate variability parameters were also comparable between groups. Low-frequency power did not differ significantly between workers exposed to chronic intermittent hypobaric hypoxia and controls (p = 0.608). High-frequency power was also similar between groups (p = 0.948), and the LF/HF ratio showed no statistically significant difference (p = 0.992) **(Fig 3)**.

**Fig 3.**
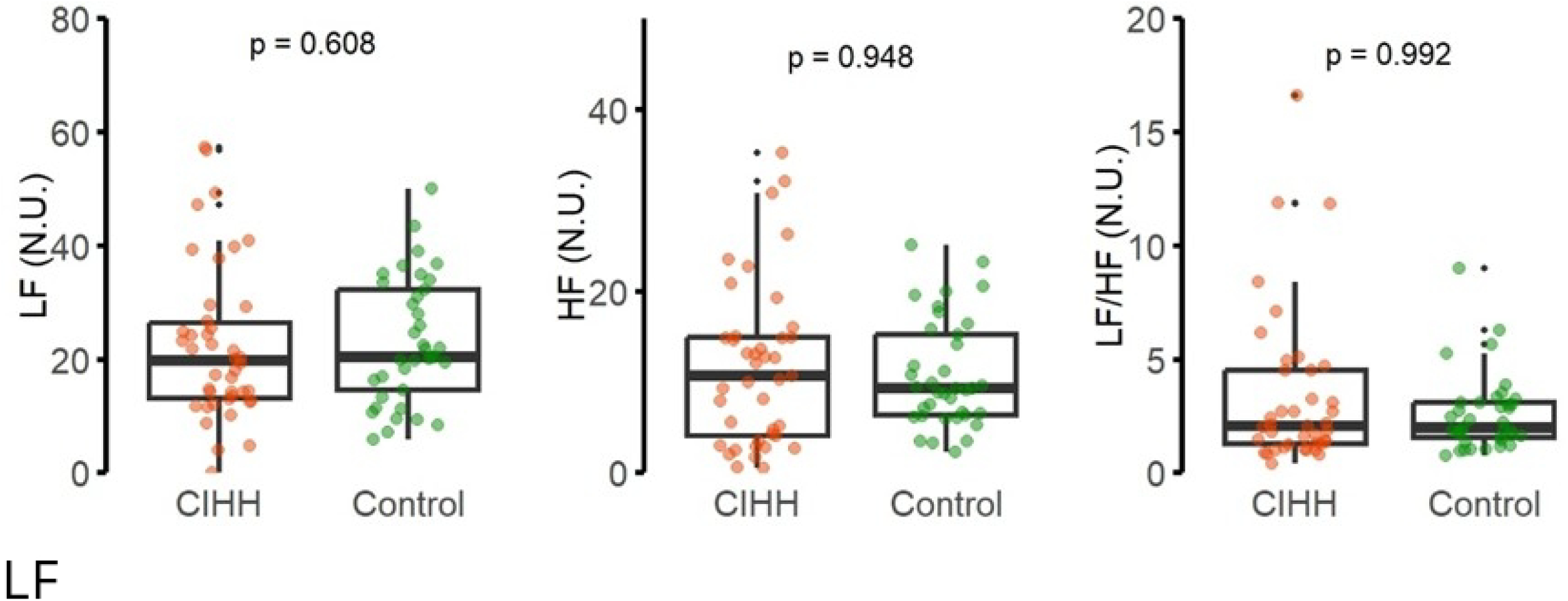
Frequency-domain heart rate variability parameters according to chronic intermittent hypobaric hypoxia exposure. The figure shows the distribution of frequency-domain heart rate variability parameters among workers exposed to chronic intermittent hypobaric hypoxia and controls. Parameters included low-frequency power, high-frequency power, and the LF/HF ratio, expressed in normalized units. Boxplots represent the median and interquartile range, with individual data points overlaid. No statistically significant differences were observed between groups. CIHH: chronic intermittent hypobaric hypoxia; LF: low frequency; HF: high frequency; N.U.: normalized units.

## Discussion

The present study compared electrocardiographic parameters and heart rate variability between workers exposed to chronic intermittent hypobaric hypoxia through rotational high-altitude work and controls working permanently at lower altitude in Arequipa, Peru. The main finding was that workers exposed to CIHH did not show statistically significant differences in PR interval, corrected QT interval, time-domain HRV parameters, or frequency-domain HRV parameters compared with controls. Vital signs, including blood pressure, heart rate, and peripheral oxygen saturation, were also clinically similar between groups. These findings suggest that, in this sample of apparently healthy male workers evaluated during their rest period at moderate altitude, CIHH exposure was not associated with measurable alterations in cardiac conduction, ventricular repolarization, or resting autonomic modulation.

HRV is widely recognized as a non-invasive marker of cardiac autonomic regulation and has been used in cardiovascular physiology, electrophysiology, occupational health, and clinical risk assessment [18,19,27,28]. Lower HRV has been associated with impaired autonomic regulation and adverse cardiovascular outcomes in different clinical settings, whereas preserved HRV is generally interpreted as a marker of more flexible autonomic control [19,27]. Therefore, evaluating HRV in workers exposed to CIHH may provide useful information on subclinical cardiovascular adaptation, even among individuals without known cardiovascular disease.

The absence of significant differences may be partly explained by the timing and setting of the measurements. Participants were evaluated in Arequipa, at approximately 2,300 m above sea level, during their rest period, rather than during active exposure at high-altitude worksites. Previous studies have shown that cardiovascular responses to intermittent high-altitude exposure may vary according to altitude level, duration of exposure, acclimatization status, work schedule, and timing of assessment [4,5,13,14]. Studies in miners and other high-altitude workers have reported changes in blood pressure regulation, cardiorespiratory function, resting metabolic rate, HRV, right ventricular parameters, and pulmonary artery pressure [11–14,29,30]. However, those findings may not be directly comparable with the present study because of differences in exposure altitude, occupational workload, cumulative exposure, study outcomes, and whether measurements were performed during active exposure, nighttime rest at altitude, or post-descent recovery.

Regarding electrocardiographic parameters, PR interval and QTc interval were similar between groups. Previous reviews have described electrocardiographic changes in populations exposed to high altitude, including right-axis deviation, repolarization abnormalities, and QTc prolongation in some settings [20–22]. However, these changes appear to depend on altitude level, speed of ascent, duration of exposure, individual susceptibility, and timing of assessment. The lack of significant differences observed in this study suggests that rotational exposure to CIHH, when assessed during rest at moderate altitude, may not produce persistent alterations in atrioventricular conduction or ventricular repolarization detectable by standard 12-lead electrocardiography in apparently healthy workers.

Time-domain HRV parameters, including SDNN, RMSSD, pNN50, and sample entropy, did not differ significantly between groups. SDNN reflects overall HRV, whereas RMSSD and pNN50 are commonly interpreted as markers more closely related to parasympathetic modulation [18,19,27,28]. Frequency-domain parameters, including LF, HF, and the LF/HF ratio, were also comparable between groups. Although LF/HF has traditionally been used as an indicator of sympathovagal balance, this interpretation requires caution because LF and HF components reflect complex interactions among autonomic, respiratory, vascular, and baroreflex mechanisms [18,19,28]. Therefore, the present findings should be interpreted as the absence of detectable differences in resting autonomic modulation under the specific measurement conditions of this study, rather than as definitive evidence that CIHH has no autonomic effect.

The methodological characteristics of HRV assessment are important for interpreting these results. Short-term HRV recordings are commonly used because they allow autonomic evaluation under controlled and feasible conditions, particularly in occupational studies [31,32]. However, HRV is sensitive to body position, environmental conditions, breathing pattern, sleep quality, recent physical activity, hydration status, psychological stress, and stimulant intake such as caffeine [31,32,33]. Although recordings in this study were obtained in the morning after a 15-minute resting period and using a device previously validated against electrocardiographic recordings [24], unmeasured pre-test factors may have contributed to within-group variability. Age and sex also influence short-term HRV, even among healthy individuals [34]. The inclusion of only male participants may have reduced biological variability but limits the generalizability of the findings to female workers exposed to CIHH.

The lack of significant differences may also reflect partial occupational adaptation. Workers exposed to CIHH repeatedly alternate between high-altitude exposure and lower-altitude recovery, which may induce adaptive or maladaptive cardiovascular responses depending on exposure intensity, duration, recovery time, and individual vulnerability [6–10]. In occupational settings, these responses may also be influenced by workload, sleep disruption, shift duration, and cumulative years of exposure [4,5,29,30]. Recent evidence from Peruvian miners exposed to intermittent hypobaric hypoxia suggests that this condition may be associated with cardiometabolic risk profiles, including obesity-related cardiovascular risk factors [35]. In this context, the present study provides a complementary perspective by showing that resting HRV and standard electrocardiographic parameters may remain comparable between CIHH-exposed workers and controls when assessed during rest-period evaluation.

This study has several strengths. It addresses an occupational exposure that is highly relevant in Andean countries, focuses on apparently healthy workers without known cardiovascular disease, uses HRV and ECG as non-invasive markers of cardiac autonomic and electrical function, and includes a comparison group working permanently at lower altitude. However, several limitations should be considered. The cross-sectional design does not allow causal inference or assessment of temporal changes. Measurements were performed during the rest period at moderate altitude rather than during active exposure at high-altitude worksites, which may have attenuated detectable differences. The sample included only male workers, and residual confounding related to work intensity, rotation schedule, sleep, diet, hydration, caffeine intake, psychological stress, breathing pattern, and recent physical activity cannot be ruled out. Finally, HRV was assessed during a single short-term resting recording, whereas repeated measurements across different phases of the work-rest cycle could provide a more complete evaluation of autonomic adaptation to CIHH.

Overall, these findings suggest that apparently healthy male workers exposed to CIHH may not present persistent differences in resting HRV or standard electrocardiographic parameters when evaluated during their rest period at moderate altitude. Future longitudinal studies should assess workers before ascent, during high-altitude exposure, during nighttime rest at altitude, immediately after descent, and after recovery to determine whether CIHH produces transient autonomic or electrocardiographic changes that are not detectable during rest-period evaluation.

## Conclusions

In this sample of apparently healthy male workers evaluated during their rest period at moderate altitude, exposure to chronic intermittent hypobaric hypoxia was not associated with statistically significant differences in resting heart rate variability or standard electrocardiographic parameters compared with controls. PR interval, corrected QT interval, time-domain HRV parameters, and frequency-domain HRV parameters were similar between groups. These findings suggest that potential autonomic or electrocardiographic effects of CIHH may not be detectable under rest-period evaluation conditions. Longitudinal studies with repeated measurements across different phases of the work-rest cycle, including active high-altitude exposure and post-descent recovery, are needed to better characterize the cardiovascular response to chronic intermittent hypobaric hypoxia in high-altitude workers.

## Data Availability

https://docs.google.com/spreadsheets/d/1OfQjkz81MZXfmnI_BevCvmMieq0ThKPF/edit?usp=sharing&ouid=108384029081479130495&rtpof=true&sd=true

## Acknowledgments

The authors thank the Vice-Rectorate of Research of the Universidad Nacional de San Agustín de Arequipa and the Project Monitoring and Support Unit for their administrative and technical support during the development of this study. The authors also thank the Faculty of Medicine of the Universidad Nacional de San Agustín de Arequipa for allowing the use of its laboratories, and Jhon Rodríguez Valdivia for his assistance in data collection. Finally, the authors thank all workers who voluntarily participated in the study.

## Data availability statement

The anonymized dataset underlying the findings of this study is available as Supporting information.

